# (How) Do Health Shocks Reallocate Research Direction?

**DOI:** 10.1101/2025.09.29.25336909

**Authors:** Hongyu Zhou, Prashant Garg, Thiemo Fetzer

## Abstract

We examine whether research systems reallocate scientific effort as health needs change. We assemble a global disease–location panel for 204 countries and territories (1990–2021) by linking disease-specific publication output to disease burden in the same place and year. Using large language models, we extract diseases from article text, map them into a standardized disease classification, and classify research funders by type. Empirically, we estimate how publication output co-moves with disease burden within countries and diseases over time, and we use event-study difference-in-differences designs that exploit plausibly exogenous variation from the timing of outbreak alerts. We find that responsiveness to endemic burden has increased over time but remains highly uneven across locations; outbreak alerts trigger rapid, statistically significant research surges that have strengthened in recent years; and funding composition is strongly associated with adjustment dynamics, with philanthropic and government-supported research contributing disproportionately to responsiveness growth in lower-income settings.

## 1 Introduction

Why do some health needs attract rapid and sustained research investment, while others remain persistently under-served? For economists, this is an allocation problem: research portfolios reflect how scientific labor, funding, and attention reallocate across topics when perceived returns or social value change, subject to frictions such as topic-specific human capital, organizational fixed costs, and path dependence. In the economics of innovation, the direction of research is known to respond to incentives, such as market size and demand-pull forces, as well as financing and time-to-payoff constraints. These forces can skew activity toward areas with stronger private returns (Acemoglu and Linn, 2004; Budish et al., 2015). In the economics of science, recent work formalizes and measures these reallocative frictions directly, showing that inducing researchers to switch topics can be costly and slow, implying limited “elasticity” of scientific direction in the short run (Myers, 2020). We ask whether the global health domain exhibits similar inertia or whether research systems actively adapt to changing health needs.

The stakes are especially high in global health because the geographic distribution of research capacity is highly unequal: although most of the world’s population lives in low- and middle-income settings, clinical research and publication leadership remain concentrated in richer systems (World Health Organization, 2024b; Lewis et al., 2021).^1^ Such spatial disparities in knowledge production shape global health equity, the external validity of clinical evidence (Bareinboim and Pearl, 2016), and the speed with which new treatments reach populations that need them most. If scientific direction responds primarily to private incentives and existing capacity, then rising burden in lower-capacity settings may translate only weakly into local research activity; conversely, if research systems are becoming more responsive, one should observe reallocation toward burden as needs evolve. Whether adjustment occurs, both gradually and abruptly, is therefore an empirical question with direct policy relevance.

Measuring adjustment is difficult. Disease burden and research output co-evolve; salience, funding, and publication incentives are jointly endogenous. Moreover, publication records and epidemiological burden data are not natively linked at comparable disease and geography levels— the world of scholarly publishing and that of disease surveillance are seldom connected by a common ontology (Røttingen et al., 2013; Mongeon and Paul-Hus, 2016; Van Leeuwen et al., 2001). Prior work documents substantial mismatches between research attention and disease burden across diseases and regions (Evans et al., 2014; Yegros-Yegros et al., 2020; Gillum et al., 2011; Access to Medicine Foundation, 2024) and highlights forces that shape the direction of science, including opportunities and expected benefits (Bhattacharya and Packalen, 2011). Yet much of this evidence is cross-sectional, aggregated, or not designed to estimate marginal responsiveness to local need. Focusing on global disease–research alignment, Schmallenbach et al. (2025) compare the distribution of publications and DALYs across diseases using a Kullback–Leibler divergence metric. They document a decline in divergence over time, but show through counterfactual decompositions that this improvement is driven primarily by shifts in disease burden rather than by active reallocation of research effort. Their analysis therefore characterizes aggregate composition rather than local responsiveness and, crucially for economic interpretation, does not pin down the elasticity of research effort with respect to need.

We address this measurement and identification problem by constructing a long-run disease-by-location panel that matches research output on each disease in each location to the corresponding burden from the same disease in the same location-year. This structure lets us estimate an “elasticity of science to health need” at the country–disease level, where resource allocation decisions and capacity constraints are plausibly organized and where frictions (e.g. specialization and cumulative advantage) should matter (Jones, 2009). It also allows us to study both slow-moving reallocations (endemic burden) and sudden re-prioritization (outbreak shocks) within a unified framework.

Our data combine publications, disease burden, outbreak alerts, and funding information at global scale. We compile 1,065,683 papers from 524 medical journals and construct a core analytic sample of 308,135 papers linked to disease and geography. For each paper we identify the diseases studied, the countries or territories whose data or patients are analysed, and the institutional homes of authors. To do so, we use large language models to extract disease and location information from titles and abstracts, then use neural embeddings to align extracted mentions to MeSH descriptors and Global Burden of Disease (GBD) categories (Global Burden of Disease Collaborative Network, 2024). We pair this with DALY-based burden series for 204 countries and territories (1990–2021), 3,134 WHO Disease Outbreak News alerts as sudden disease-shock events, and a funder classification spanning public, corporate, philanthropic, academic, and hybrid types.

We report three main findings. First, endemic responsiveness has risen substantially: research output has become more elastic to domestic disease burden over time, though large cross-location disparities persist. Second, emergency responsiveness is sizable and increasingly strong: in an event-study difference-in-differences design with country–disease, country–year, and disease– year fixed effects, together with estimators robust to staggered timing (Sun and Abraham, 2021; Callaway and Sant’Anna, 2021; Goodman-Bacon, 2021), outbreak alerts trigger rapid and statistically significant increases in disease-specific research. Third, funding composition is strongly associated with adjustment dynamics: philanthropic foundations disproportionately support research into diseases concentrated in lower-income settings, whereas corporate R&D tilts toward diseases associated with higher stages of economic development; in lower-income settings, a substantial share of responsiveness growth is linked to philanthropic and government-supported research. Taken together, results paint a cautiously optimistic picture, yet significant misalignments endure.^2^

Conceptually, our findings speak to marginal responsiveness at the country–disease level, distinct from aggregate compositional alignment. This distinction is policy-relevant because research funding, infrastructure, and agenda setting are largely organized at the national level, and because reallocating scientific effort may be constrained by specialization and cumulative advantage. Our framework therefore permits assessment of whether research systems dynamically adapt to local disease burden, how quickly they respond to sudden shocks, and where capacity is expanding or lagging.

Our contribution is threefold. First, we contribute to the economics of science and innovation by estimating need-based elasticities of research effort and by quantifying shock-driven re-prioritization, complementing work on the direction and elasticity of science and on how opportunities and incentives shape research direction (Myers, 2020; Bhattacharya and Packalen, 2011; Acemoglu and Linn, 2004; Budish et al., 2015). This framing also connects to metascience on (mis)alignment and shock-driven reallocation (Gillum et al., 2011; Evans et al., 2014; Yegros-Yegros et al., 2020; Furuse, 2019; Ioannidis et al., 2022; Wagner et al., 2022; Zhao et al., 2022; Asgari et al., 2025) by shifting attention from descriptive mismatches toward estimable responsiveness parameters.

Second, we contribute to work on global health research (mis)alignment and the geography of science by moving beyond aggregate composition metrics to identify within-country, within-disease responsiveness—the margin at which national systems can reallocate even when global shares remain stable (Schmallenbach et al., 2025). In doing so, we also speak to science studies on geography, collaboration, and cumulative advantage (Merton, 1968; Hoekman et al., 2010; Adams, 2012; Fortunato et al., 2018; Castro Torres and Alburez-Gutierrez, 2022; Dajani, 2023) and to concerns about external validity and where evidence is produced relative to where burden lies (Bareinboim and Pearl, 2016).

Third, methodologically, we provide a scalable extraction-and-alignment pipeline that links publications to disease and place over long horizons, enabling disease-by-location measurement at global scale and supporting decomposition exercises that attribute responsiveness growth to portfolio composition and funding institutions (Azoulay et al., 2019; Chu and Evans, 2021; Sattari et al., 2022).^3^ Empirically, we combine endemic burden gradients with outbreak-based event studies, allowing us to study both slow-moving and sudden drivers of research reallocation within a unified framework.

The remainder of the paper proceeds as follows. The data section describes data construction and measurement. The empirical strategy section sets out identification for endemic burden responsiveness, outbreak-shock event studies, and funder decomposition. The results and discussion section presents the main findings and robustness analyses. The conclusion section closes.

## 2 Data and Measurement

We construct a disease–location–year panel that links publication output to disease burden and outbreak shocks using an LLM-assisted extraction-and-normalisation pipeline. Figure 1 summarizes the end-to-end workflow: (i) selecting a medical-journal corpus in OpenAlex, (ii) extracting diseases and study locations from titles/abstracts, (iii) embedding-based normalisation to MeSH and a crosswalk to GBD causes, and (iv) merging to DALYs and WHO outbreak alerts for the endemic and event-study designs.

**Figure 1:**
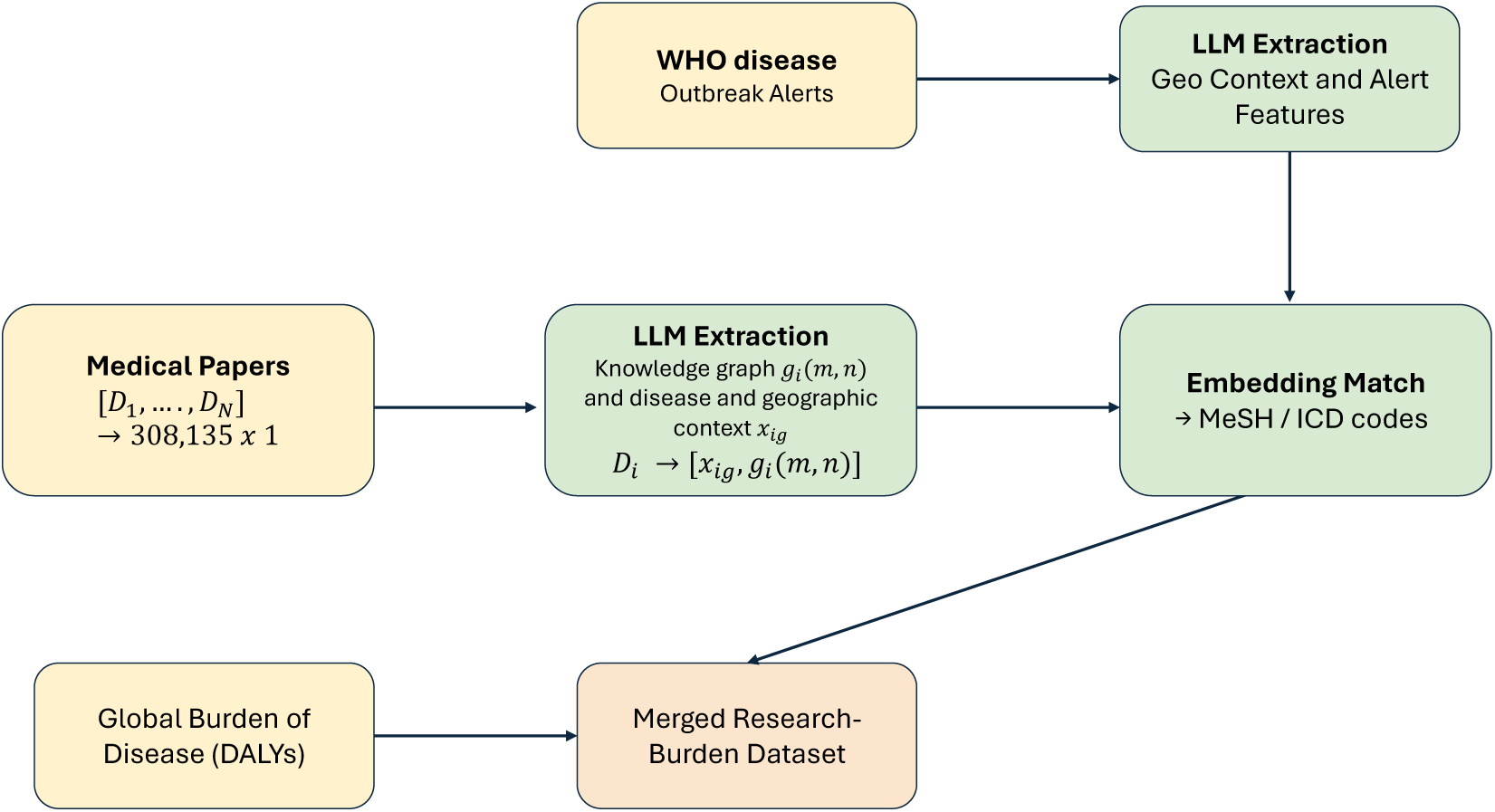
Overview of the data construction and linking pipeline. **Notes:** Starting from 308,135 medical papers in OpenAlex, we use LLMs to extract disease entities and geographic context from titles/abstracts. Extracted disease strings are embedded and matched to MeSH descriptors (and crosswalked to ICD/GBD codes), enabling a merge to location-year DALYs from the Global Burden of Disease to form the research–burden panel. In parallel, WHO Disease Outbreak News alerts are processed with an LLM to extract disease, geography, and alert features, which are normalized to the same ontology and merged to the panel for event-study estimation.

### 2.1 Paper–Disease–Location Panel

#### Publication corpus and sample construction

Publication records come from OpenAlex (Priem et al., 2022). We start from 1,065,683 papers in 524 medical journals drawn from the 2023 *Journal Citation Reports* categories *Medicine, General & Internal*, *Medicine, Research & Experimental*, and *Pharmacology & Pharmacy*. We restrict to English-language Health Sciences research articles with citation and affiliation metadata and retain papers with potentially identifiable geographic content in titles/abstracts using complementary location-detection approaches (a named-entity-recognition model such as distilbert-NER and the GeoText dictionary). This yields a core corpus of 308,135 papers used for LLM extraction (Appendix Section A).

#### LLM extraction of diseases and study locations

For each paper, we use a single LLM call on title and abstract to extract (i) disease strings and (ii) study-location country mentions (ISO-3 codes) in a structured JSON format (Appendix Section B). The extraction prompt is designed to be conservative: it records only entities explicitly mentioned and defaults to empty fields when information is absent, reducing hallucination risk. We use fractional counting when a paper mentions multiple diseases and/or locations, so that each paper contributes at most one unit of output distributed across its extracted disease–location combinations.

We use gpt-4o-mini to extract entities from titles and abstracts, returning a JSON object under a strict schema with fixed keys and disallowed additional fields (Appendix Section B). This structured-output setup enforces parseability and reduces hallucinated fields: missing information is recorded as empty arrays or NA rather than inferred. We treat extraction as mention-level rather than classification: the model is instructed to record only diseases and locations explicitly stated in the input text. We cache model outputs and retain the raw JSON for auditing and replication.

#### Embedding-based normalisation to MeSH and mapping to GBD

Extracted disease strings are normalised to a controlled biomedical vocabulary by embedding each string and matching it to the nearest MeSH descriptor (National Library of Medicine, 2023; Sampson et al., 2009). We then map MeSH descriptors to Global Burden of Disease (GBD) causes using a MeSH–GBD crosswalk, allowing direct linkage between research output and disease burden. Using this pipeline, approximately 86% of papers map to level-2 GBD causes; certain rare pathogens (e.g. MERS coronavirus) lack direct GBD codes and are excluded from burden-adjusted regressions but retained for WHO-alert analyses.

#### Deterministic normalisation and multi-entity handling

Normalisation is fully deterministic conditional on the extracted strings. Each extracted disease string is embedded using text-embedding-3-large and matched to the nearest MeSH preferred descriptor among 30,836 candidates by cosine similarity (Appendix Section B.2). We validate this normalisation against PubMed’s human-assigned MeSH annotations and find agreement of 92.9% at GBD level 2 and 91.2% at GBD level 3 (Appendix Section B). When a paper mentions multiple diseases or locations, we construct all implied disease–location pairs and use fractional counting so that each paper contributes a total weight of one across its extracted pairs.

#### Disease burden linkage and panel unit

We merge publication output to DALY-based burden series from the GBD dataset for 204 countries and territories over 1990–2021 (Vos et al., 2020; Global Burden of Disease Collaborative Network, 2024). The resulting analytic dataset is organised at the disease–location–year level, with annual publication output defined as the (fractionally counted) number of papers that mention disease *i* and study location *ℓ* in year *t*, and burden measured as DALY*_iℓt_* for the corresponding disease and location-year (Figure 1).

#### Funding data and funder-type classification

OpenAlex provides funding acknowledgements for a subset of papers; we use these to study how portfolio composition differs by funder type and how funders contribute to responsiveness growth. We classify each unique funding institution into public, corporate, philanthropic, academic, and hybrid categories using a multi-round LLM protocol: each funder name is classified in five repeated runs and assigned its modal label, yielding high internal agreement (96–99%). Full prompt and aggregation rules are provided in Appendix Section D.

### 2.2 Disease-Shock Data

To capture sudden health shocks, we assemble WHO Disease Outbreak News alerts from 1996 to 2025. The resulting dataset contains 3,134 alerts on 2,481 dates, each with event text and publication date (see Appendix Section C for full details of our information extraction prompt). We parse alert text to recover disease–geography–time mentions and map disease mentions to the same MeSH/GBD ontology used in the main panel. This produces a harmonized event dataset that can be merged to disease-by-location outcomes for event-study estimation.

Because alert text often mixes coarse and fine geographic references, we standardize location mentions and assign deterministic identifiers before merging. For records with incomplete time mentions, we use the alert publication date to recover event year. When multiple alerts occur for the same disease–location cell within a year, we keep the first alert date as the event-time anchor and treat subsequent alerts as part of the same annual shock episode.

### 2.3 Predictors for Response Heterogeneity

For heterogeneity analysis, we construct a predictor set spanning research capacity, demographics, digital connectivity, macroeconomic conditions, governance, health-system preparedness, disease burden, and text-derived alert features. These variables are assembled as covariates for a secondary prediction exercise and are not used as primary identification devices in the baseline econometric designs. We align predictors to pre-event or contemporaneous information where applicable; the exact response targets and model setup are introduced in Section 3. Table 1 summarizes predictor categories, representative variables, constructions, and primary data sources used in the heterogeneity analysis.

**Table 1:** Predictor summary for heterogeneity analysis.

| Variable | Definition / construction |
| --- | --- |
| H index | Country-level bibliometric proxy of cumulative research impact. |
| Average citations | Mean citations per publication at country level. |
| Citable documents | Country-level volume of citable scientific output. |
| Population | Total country population by year. |
| Urban share | Share of population living in urban areas. |
| Population density | Population per square kilometer of land area. |
| Share age 65+ | Share of population aged 65 and above. |
| Internet use share | Share of population using the internet. |
| GDP per capita | Country-year income level per person. |
| Government effectiveness | Institutional effectiveness and policy implementation capacity. |
| Political-security risk | Indicator of political and security vulnerability. |
| Socio-economic resilience | Capacity to absorb and recover from shocks. |
| Public confidence in government | Indicator of trust in government institutions. |
| HAQ index | Healthcare access and quality proxy. |
| GHS pillars | Preparedness domains for epidemic prevention, detection, and response. |
| DPT3 coverage | Immunization coverage among children ages 12–23 months. |
| Mean DALY rate (alerted disease) | Disease burden in affected countries; one-to-many mappings are averaged across mapped causes. |
| Alert length | Token/character-length proxy for report detail and salience. |
| Uncertainty lexicon features | Dictionary-based counts for overall uncertainty and subtypes (hedging, possibility, confusion). |

In sum, our empirical dataset is organized at the disease–location–year level. This structure supports two complementary designs used below: endemic responsiveness from within-location, within-disease burden variation over time, and emergency responsiveness from event-time variation around outbreak alerts. Detailed variable definitions, source tables, matching rules, transformation choices, Random Forest inputs, and additional robustness checks are provided in the appendix; see Appendix Sections A–C.

## 3 Empirical Strategy

### 3.1 Estimating Research–Disease Elasticity (Endemic Responsiveness)

We estimate how research output responds to gradual changes in disease burden by fitting fixed-effects panel models at the disease–location–year level:

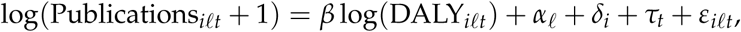

where *i* indexes diseases, *ℓ* locations, and *t* years. The elasticity parameter *β* captures the percentage change in research output associated with a one-percent change in burden for the same disease in the same location. Identification comes from within-panel variation after conditioning on location fixed effects (*α_ℓ_*), disease fixed effects (*δ_i_*), and year fixed effects (*τ_t_*), which absorb time-invariant cross-location differences, persistent disease-specific publication levels, and common global time shocks.

We implement three complementary endemic specifications. First, the baseline elasticity model above provides the average burden responsiveness across the full panel. Second, to characterize temporal and distributional dynamics, we interact log(DALY*_iℓt_*) with year and income-group indicators and summarize year–income-group-specific elasticities. This allows us to assess whether responsiveness rises uniformly over time or diverges across income strata. Third, to capture cross-country heterogeneity, we estimate country-specific burden elasticities by interacting log(DALY*_iℓt_*) with location indicators.

These specifications map directly to the endemic results we report: aggregate responsiveness, dynamic income-group trajectories, and cross-country heterogeneity. We then use the resulting elasticity estimates as inputs to decomposition exercises that attribute changes in responsiveness to disease composition and funding composition. Standard errors are clustered at the disease– location level to account for serial correlation within panel cells.

### 3.2 Event-Study Design for Outbreak-Shock Responsiveness

To identify research responses to sudden health shocks, we implement an event-study difference-in-differences design using WHO Disease Outbreak News alerts as treatment events. We treat a WHO alert on a specific disease in a particular country and year as an exogenous shock—an unexpected event that raises the salience and perceived urgency of the disease in that context— and compare research activity before and after the alert, as well as between alerted and non-alerted disease–location pairs, to estimate how much scientific attention responds to sudden public health emergencies. The treatment is defined at the disease–location–time level: an alert for disease *i* in location *ℓ* at date *t_k_*. Our approach exploits variation in the timing and targeting of alerts across diseases and countries; by combining temporal contrasts (before vs. after the alert) with cross-sectional contrasts (treated vs. control pairs), we aim to distinguish the impact of alerts from underlying secular trends or shared shocks. The fixed-effects structure below is designed to absorb confounders at the country, disease, and global level so that the estimated coefficients can be interpreted as shifts in research activity following alerts rather than artifacts of pre-existing trends.

Let *k* index alert events and *τ* denote event time:

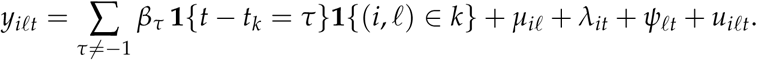

Here, *y_i__ℓt_* is annual disease–location publication output, measured as log(Publications*_iℓt_* + 1), and *τ* = −1 is omitted as the reference period. We estimate dynamic coefficients over a symmetric five-year pre/post window around each alert (*τ* ∈ {−5, …, +5} \ {−1}). The coefficients *β_τ_* trace responses before and after alerts and are interpreted as event-time treatment effects relative to the period immediately preceding the alert.

A central challenge is counterfactual construction. We primarily use a between-disease, between-country comparison, benchmarking alerted disease–location pairs against all other disease– location combinations. This provides broad identifying variation and a transparent baseline for interpreting event-time coefficients. To assess whether findings depend on control-group definition, we also estimate two alternative designs as robustness checks: (i) within-disease, between-country comparisons, where treated units are compared with the same disease in non-alerted locations; and (ii) between-disease, within-country comparisons, where treated disease–location pairs are compared with other diseases in the same location-year.

The fixed-effects structure is designed to absorb major confounders at multiple margins. Country–disease fixed effects (*µ_iℓ_*) absorb persistent differences in baseline research intensity across disease–location pairs. Disease–year fixed effects (*λ_it_*) absorb global disease-specific shocks, such as worldwide scientific or policy attention shifts. Location–year fixed effects (*ψ_ℓt_*) absorb location-wide annual shocks, including broad changes in research capacity, funding, or policy environments. Together, these controls isolate within-cell deviations around alert timing from broader secular trends.

Identification relies on the absence of differential pre-trends between treated and control units after conditioning on these fixed effects, which we assess directly from pre-alert event-time coefficients. Standard errors are clustered at the disease–location level to allow for serial correlation within panel cells.

We report both pooled and dynamic summaries. Pooled estimates collapse post-alert periods into an average effect for concise cross-context comparison; dynamic estimates retain the full event-time profile to assess response timing, persistence, and pre-trends. Results from alternative comparison-group specifications and additional robustness analyses are reported in the appendix.

### 3.3 Decomposing Funder Impacts on Elasticity Change

Which funder types contribute most to the growth of endemic responsiveness? To answer this, we decompose changes in burden elasticity across periods by funding source. For each funder type *f*, we construct a counterfactual panel that removes publications supported by *f*, re-estimate elasticity growth under the same endemic specification, and compare it with baseline growth. The difference provides an attribution measure of each funder type’s contribution to observed responsiveness gains, including whether contributions differ across income groups.

In parallel, we characterize portfolio orientation by funder type using a funding-tilt statistic, so that contribution estimates can be interpreted jointly with differences in funded disease mix.

For funder type *f* and disease group *d*, we define:

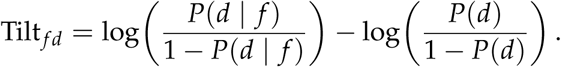

These decomposition estimates are attributional summaries of portfolio-linked responsiveness changes, rather than causal treatment effects of funder types. Removing a funder type reduces estimated elasticity growth when the papers it supports are disproportionately concentrated in disease–location cells where burden-driven research has expanded; the contribution measure thus attributes observed responsiveness gains to the portfolio orientation of each funder, without implying that funder type causes responsiveness.

### 3.4 Explaining Heterogeneity in Response Magnitudes Using Random Forest Regression

Finally, we use Random Forest models to explain cross-context variation in estimated response magnitudes from the event-study stage. Our primary prediction target is the pooled estimate, and we train models using a broad set of indicators spanning research capacity, demographics, digital connectivity, macroeconomic conditions, governance, health-system preparedness, disease burden, and alert-text characteristics. This setup allows us to summarize which observable country-, disease-, and alert-level factors are most informative about where emergency responsiveness is larger or smaller.

Beyond pooled effects, we also examine a dynamic-response target based on Δ*β*, which captures changes in event-time coefficients across post-alert horizons. This complementary exercise assesses whether predictors of average responsiveness are the same as predictors of response dynamics. We report feature-importance results for the pooled target in the main article, while Δ*β*-target results are reported in the appendix. In all cases, these machine-learning analyses are descriptive complements to the main econometric identification strategy, and full predictor definitions, model configurations, and robustness results are reported in the appendix.

## 4 Results and Discussion

### 4.1 Disparities and shifts in global disease research

We begin by charting the global landscape and evolution of disease research in relation to disease burden. Across our dataset of research papers, the majority of work focuses on cardiovascular diseases and neoplasms, while comparatively less attention is directed toward neglected tropical diseases, malaria, and nutritional deficiencies (Figure 2a). However, the volume of research efforts does not correspond closely to patterns of disease burden (disability-adjusted life years, DALYs) globally. For instance, respiratory infections and tuberculosis, as well as maternal and neonatal conditions, exhibit a much higher disease burden relative to their paper count ranking, whereas neoplasms, digestive diseases, and diabetes and kidney diseases are overrepresented in research output relative to their burden. On the other hand, emergency disease outbreaks, measured by WHO alerts, are most frequently associated with infectious diseases, particularly respiratory infections, tuberculosis, and neglected tropical diseases and malaria.

**Figure 2:**
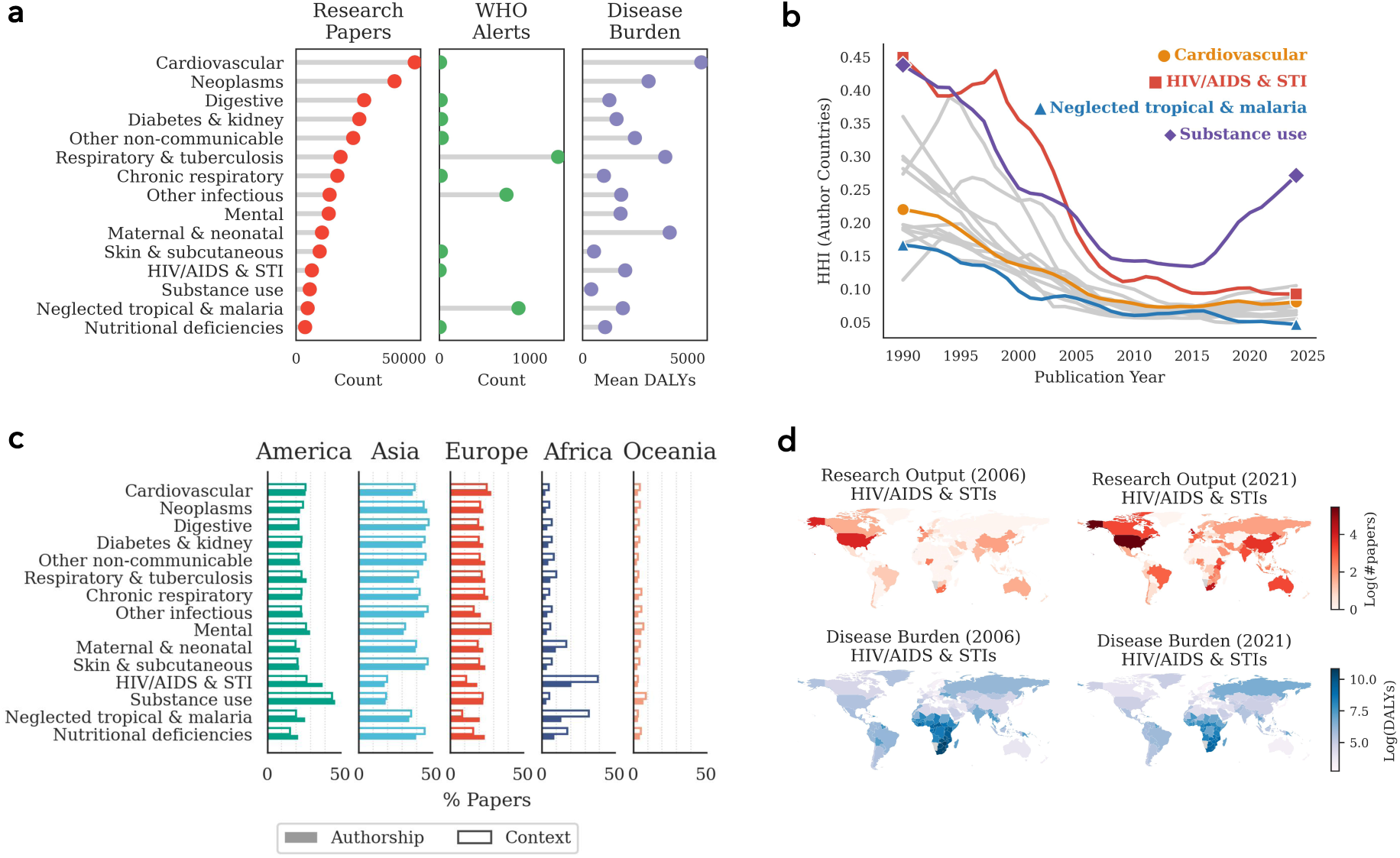
Global representation and evolution of disease research and burden. **Notes: a,** Cross-comparison of fifteen disease groups by total number of research papers, number of WHO alerts, and mean disability-adjusted life years (DALYs). **b,** Temporal trends in the geographic concentration of research, measured by the Herfindahl-Hirschman Index (HHI) of author countries. A general decline indicates increasing diversity of national contributions across most disease groups. Lines show LOWESS-smoothed trajectories. **c,** Continental share of research output (authored articles) and research context (in-text mentions) by disease group, calculated using fractional counting. **d,** Illustration of increasing alignment between research effort and disease burden for HIV/AIDS and STIs in 2006 and 2021. Research output (upper) and disease burden (lower) are shown on log-transformed scales.

A geographic lens further highlights disparities. As expected, the Americas, Asia, and Europe account for the largest shares of research authorship and in-text mentions, while Africa lags behind (Figure 2c). Yet the imbalance is nuanced. For neglected tropical diseases and malaria, Africa accounts for 32.5% of research context but only 13.9% of authorship (two-proportion *z*-test_(context, authorship)_: *z* = −20.3, *p <* 0.001). This imbalance means that regions often feature prominently as objects of study but less as sources of authorship. The pattern recurs across disease groups in Africa.

Encouragingly, over time we observe decreasing geographic concentration of research efforts (Figure 2b, see appendix), as measured by the Herfindahl–Hirschman Index (HHI), suggesting that research across most disease groups has become more geographically diverse. An exception emerges in substance use disorders, where research concentration has increased since 2015, largely reflecting the growing dominance of contributions from the United States (see appendix).

But does such geographic diversification also mean that research efforts are increasingly directed toward the regions bearing the greatest burden, thereby leading to greater responsiveness globally? Figure 2d provides an illustrative example in the case of HIV/AIDS and STIs. In 2006, the burden fell disproportionately on the Global South, particularly sub-Saharan Africa, yet research efforts from the region were limited. By 2021, while the disease burden remained highest, several countries in sub-Saharan Africa had emerged as leading contributors. This example hints at increasing responsiveness, yet unevenly, prompting us to examine the pattern more formally and empirically across countries and diseases.

### 4.2 Research responsiveness to endemic disease burden

We conceptualize endemic research responsiveness as the extent to which countries adjust their research output in line with changes in their endemic disease burden, reflecting how they structure their research portfolios and set priorities amid shifting health challenges. To capture this empirically, we estimate fixed-effects regressions of national publication counts on DALYs, controlling for country, disease, and year. The resulting coefficient, which we term elasticity, measures the percentage change in publications associated with a 1% change in DALYs (see Section 3). This framework allows us to examine how responsiveness varies across countries, diseases, and over time.

Similar to other areas of national capacity, marked disparities exist in endemic research responsiveness. Responsiveness tends to rise with GDP per capita (Pearson’s *r* = 0.43, *p <* 0.001, *R*^2^ = 0.18; Figure 3a), but high-income countries vary widely: nations such as Japan, Saudi Arabia, and the United States lie above the trend, whereas Luxembourg, Switzerland, and Iceland fall below it. Comparable disparities also emerge across diseases. Estimated as deviations from burden-expected levels, conditions prevalent in high-income settings, such as cardiovascular and digestive diseases, receive on average about 16.5% (95% CI 15.0–18.1%) and 14.1% (95% CI 12.8–15.5%) more publications than expected, respectively, whereas nutritional deficiencies and maternal and neonatal disorders attract roughly 14.4% (95% CI 13.4–15.5%) and 12.4% (95% CI 11.4–13.5%) fewer (all *p <* 0.001; Figure 3c, see appendix).

**Figure 3:**
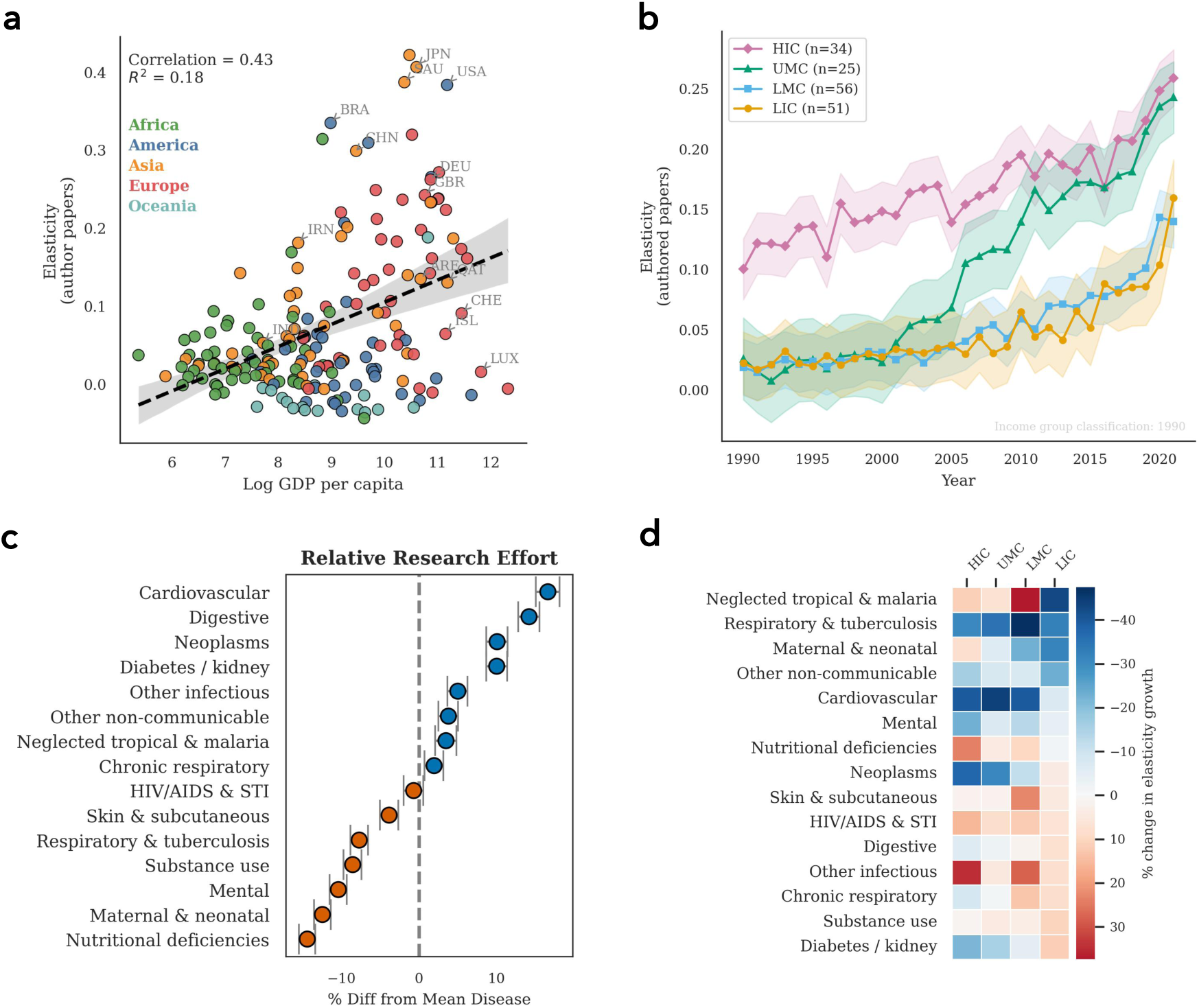
Endemic responsiveness of national research efforts to disease burden. **Notes: a,** National endemic research responsiveness positively correlates with economic development status: elasticity rises with log GDP per capita, although wealthier countries display greater heterogeneity. Each point denotes a country or territory (coloured by continent); the dashed line is the OLS fit with 95% CI. Elasticities are estimated from regressions of log publications on log DALYs with country, disease, and year fixed effects. **b,** Responsiveness has increased over time across all income groups, with high-income countries consistently exhibiting higher elasticity. Lines show estimated elasticities from regressions with year × income-group interactions; shaded areas denote 95% confidence intervals. **c**, Relative research effort across disease groups, measured as baseline deviations from burden-expected levels derived from disease fixed effects (positive/blue = over-represented relative to burden; negative/orange = under-represented). **d,** Disease-level contributions to endemic responsiveness growth between 2014–2017 and 2018–2021, by income group. For each disease, we construct a counterfactual by holding its post-2017 publication counts constant at 2017 levels and re-estimating elasticity. The heatmap shows the percentage difference from the baseline change, indicating whether growth in that disease’s research amplifies (red) or dampens (blue) the overall increase in responsiveness of the income group.

Building on our earlier observation of HIV/STI and AIDS, we observe a clear and sustained increase in responsiveness over time, estimated from fixed-effects regressions with year–income-group interactions. However, the pace of growth differs sharply across income groups: responsiveness roughly doubled in high-income countries between 1990 and 2021, nearly tripled in upper-middle-income countries, and rose more modestly in lower-middle- and low-income groups, as shown in Figure 3b (also see appendix). This rise offers grounds for cautious optimism, yet substantial gaps persist, with high- and upper-middle-income countries consistently leading while lower-income groups remain well behind.

These gains are not driven by research on all diseases equally. To further unpack which diseases contribute most to rising responsiveness within each income group, we construct a counterfactual that holds post-2017 publication counts constant at 2017 levels and then re-estimate the change in elasticity. This approach allows us to isolate the contribution of specific diseases to overall growth in responsiveness. The decomposition shows that the increase in responsiveness among low-income countries is amplified mainly by more research on neglected tropical diseases and malaria, respiratory infections and tuberculosis, and maternal and neonatal conditions (Figure 3d). By contrast, in high-income countries, the gains come primarily from expanded research on cardiovascular diseases and neoplasms, a pattern also observed in upper-middle-income countries. Overall, this shows that increasing endemic research responsiveness stems from more research on the conditions most responsible for disease burden in each income setting.

### 4.3 The portfolio preference and impact of research funders

Funding agencies play a central role in shaping the direction and focus of scientific research, an influence that is especially consequential for the Global South, where external resources often determine whether local health challenges receive adequate attention. To examine how funders shape disease research and contribute to responsiveness growth, we classified all 32,437 funders listed in the 2024 OpenAlex snapshot into six categories: direct government, independent public, private corporate, private philanthropic, academic institution, and hybrid, and study their differences in both funding tilt across diseases and their impact on responsiveness growth.

To examine how funders allocate resources across diseases, we calculate each type’s funding tilt, defined as the log-odds ratio of a disease being funded by that type relative to the overall average. Funding tilt varies sharply across funder types, highlighting their divergent funded disease research portfolios (Figure 4a). Private philanthropic funders tilt strongly toward nutritional deficiencies as well as HIV/AIDS and neglected tropical diseases, which disproportionately affect low-income countries. By contrast, corporate funders allocate relatively little to these conditions and instead show a strong orientation toward non-communicable diseases, particularly cardiovascular diseases, neoplasms, and diabetes and kidney diseases, which dominate in high-income settings.

**Figure 4:**
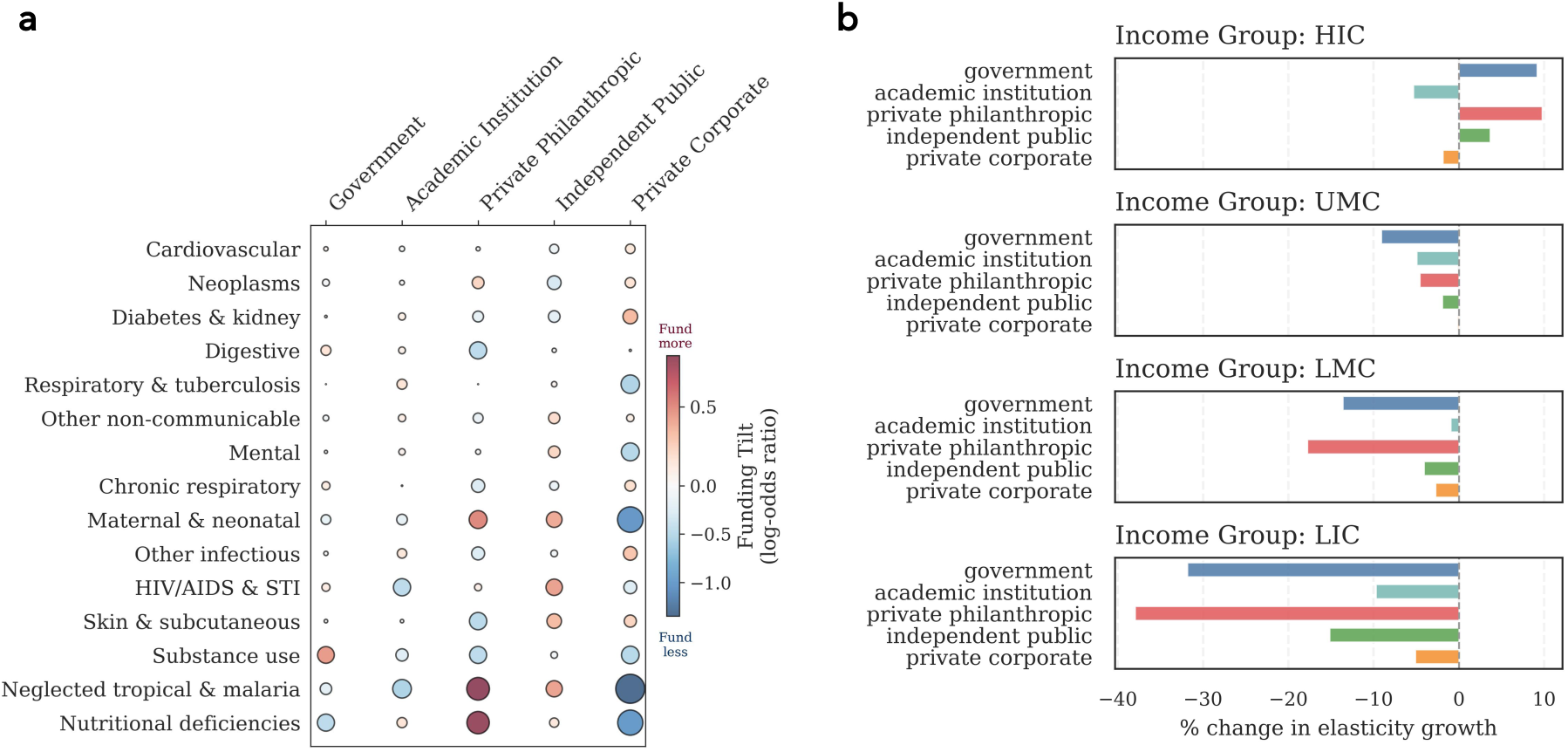
The preference and impact of research funders on disease research and endemic responsiveness. **Notes: a,** Funding tilt of five funder types across disease groups. Tilt is measured as the log-odds ratio of a disease being funded by a given type compared to the overall average. Positive values (red) indicate diseases over-represented in a funder’s portfolio, while negative values (blue) indicate under-representation. Bubble size denotes the magnitude of the tilt. **b,** Contribution of funder types to changes in research responsiveness (elasticity growth) between 2014–2017 and 2018–2021, by income group. For each funder type, we construct a counterfactual by removing all papers funded by that type and re-estimating elasticity growth. Bars show the percentage difference relative to baseline growth for each income group. Positive values indicate that a funder type makes a net positive contribution to responsiveness growth in that group (removing it reduces growth), while negative values indicate that removing the funder increases growth.

The consequences of these portfolio choices are also visible in their contributions to endemic responsiveness growth (Figure 4b). Similar to our earlier counterfactual analysis, we estimate each funder type’s impact by assuming that papers it supported were absent and re-calculating how elasticity changed between 2014–2017 and 2018–2021. We observe that, in low-income countries, responsiveness gains rely heavily on philanthropic and government support: without papers funded by them, the estimated responsiveness growth would shrink by 38.0% and 31.8%, respectively. Similar patterns are observed in lower-middle-income countries. In high-income countries, however, the pattern reverses: removing papers funded by governments or philanthropic organizations significantly reduces responsiveness growth, i.e., funding research on conditions not most prevalent in their own settings but aligned with a broader global health agenda. Put differently, these two funder types play a central role in sustaining research that extends beyond conditions prevalent in high-income settings, by directing attention toward diseases that impose heavier burdens in low-income contexts.

### 4.4 Research responsiveness in emergency: event-study evidence from WHO disease outbreak news

Outbreaks create another form of disease burden, producing sudden shocks that often leave countries unprepared. To assess how research systems respond under such conditions, we focus on emergency responsiveness, which is the capacity to scale up research activity in the wake of acute health shocks. We study this using nearly three thousand outbreaks announced by the WHO. For each outbreak, we estimate a difference-in-differences event-study specification that compares changes in research activity on the focal disease in the affected country before and after the outbreak to contemporaneous changes in other diseases and other countries. The model includes country–disease fixed effects to absorb persistent heterogeneity in research portfolios, disease–year effects to capture global shocks to individual disease areas, and country–year effects to account for overall shifts in national output. The resulting estimates capture the increase in the likelihood of publication on the focal disease after an outbreak, reflecting the capacity of national research systems to mobilize and deliver science in response to health emergencies.

Tracing evolution over time, Figure 5a shows once again increasing emergency research responsiveness: outbreak-triggered research surges were minimal in the 1990s and modest in the 2000s, but became pronounced and statistically robust in the 2010s, with outbreaks generating substantial and near-immediate increases in research output activity (see appendix for results with alternative event-study design and robustness checks). This pattern suggests that research effort allocation may increasingly anticipate disease risks. We show the robustness of this finding to a broad menu of modeling choices in the appendix by documenting a generally increasing trend in before–after shifts in event-study coefficients across years and specifications.

**Figure 5:**
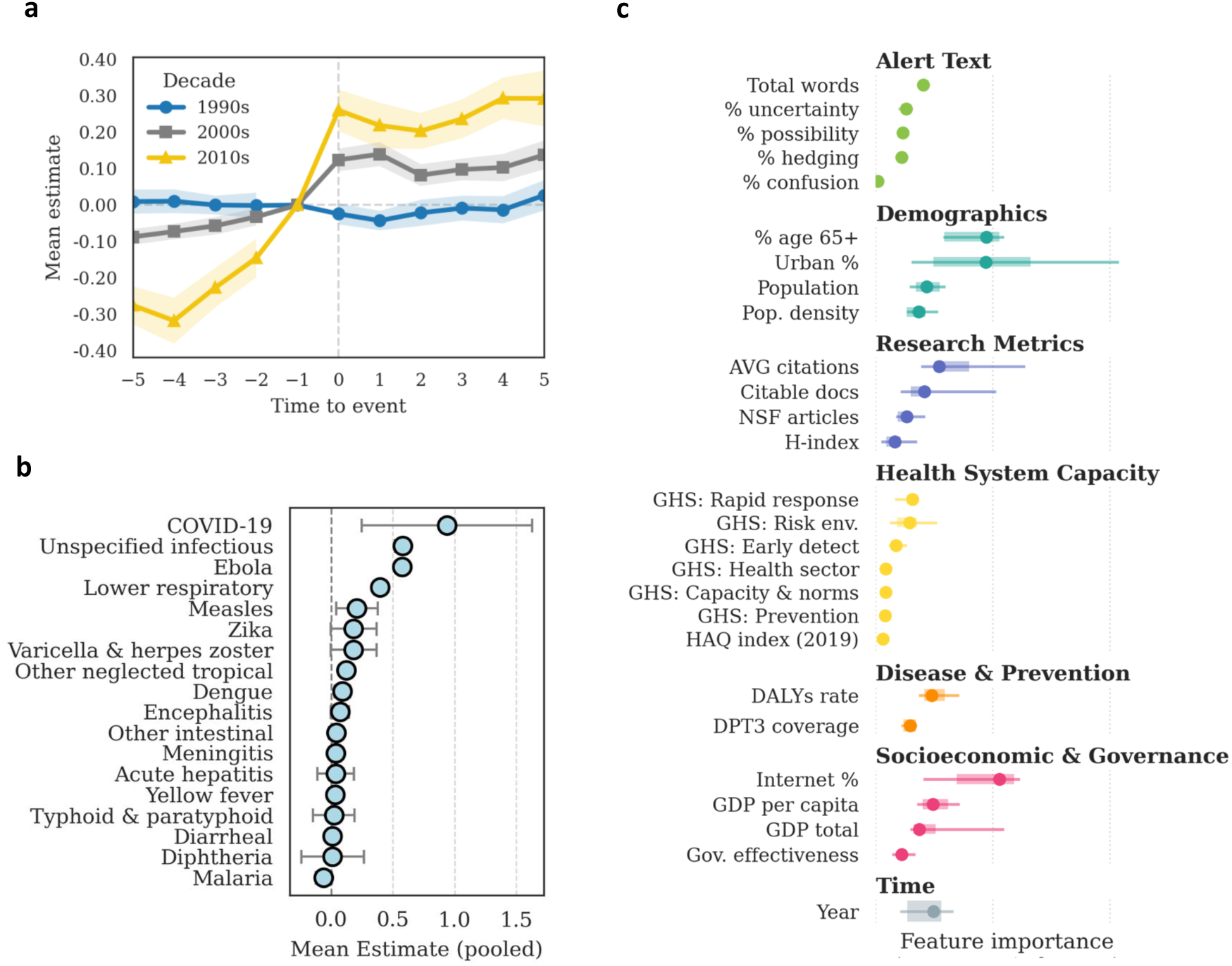
Event–study estimates and predictors of emergency research responsiveness. **Notes:** We implement an event-study difference-in-differences design for each alert–country pair to measure the growth of research on the focal disease relative to control groups. The figure shows results from the between-disease, between-country specification, which benchmarks alerted disease–country pairs against all other disease–country combinations. Alternative designs (between-disease within-country and within-disease within-country) are reported in the appendix. The estimation yields two types of coefficients: pooled estimates, summarizing the average postevent increase in research activity, and dynamic estimates, tracing the yearly evolution of responsiveness around the outbreak. **a,** Temporal change in mean responsiveness by decade. Shaded areas denote 95% confidence intervals (same for panels b and c). **b,** Average pooled estimate across disease groups (GBD level 3). **c,** Feature importance from Random Forest regressors predicting pooled estimates of responsiveness.

Not all disease alerts, however, trigger substantial research responses. Figure 5b reports the mean pooled estimate across alerts and highlights considerable variation. Emerging and high-profile pathogens such as COVID-19, Ebola, Zika, and measles generated the largest increases in related research activity, whereas endemic or historically persistent conditions such as malaria, diphtheria, and diarrheal diseases elicited weaker responses (also see appendix). These patterns suggest that research systems tend to mobilize disproportionately around novel or highly salient threats, where existing knowledge is limited and rapid scientific exploration is required.

Which factors explain variation in emergency research responsiveness? To answer this, we employed Random Forest regression models to predict the size of the estimated research response (pooled estimate) using a broad set of country-, disease-, and alert-level features (see Section 3). Figure 5c shows that four predictors stand out: population structure (share of senior citizens and urban residents), internet penetration, research capacity (average citations per publication), and the length of outbreak alerts. These results indicate that demographic structure, digital infrastructure, research preparedness, and the nature of alerts are important factors in explaining a nation’s ability to mobilize research during health crises (see appendix).

## 5 Conclusion

This paper asks whether research systems dynamically reallocate effort as health needs evolve. Our results quantify the responsiveness of knowledge production to changes in health need, and highlight how frictions in specialization, funding, and research capacity shape the speed and incidence of adjustment. Our burden-responsiveness estimates provide a quantitative complement to prior calls to align research portfolios with health need (Røttingen et al., 2013) and to address structural inequities in who leads and benefits from global health science (Hedt-Gauthier et al., 2019; Abimbola et al., 2021). Across a global disease–location panel, we find rising responsiveness on two margins: research output has become more elastic to domestic endemic burden, and outbreak alerts trigger faster and stronger disease-specific research surges than in earlier decades. The upward trend is encouraging and suggests that efforts to expand research capacity through open-access platforms, regional training, and cross-border collaborations may be bearing fruit. At the same time, adjustment remains uneven across income groups and disease areas, consistent with persistent capacity constraints and cumulative advantage in where scientific inputs are located.

Funding composition is a central part of this pattern. In lower-income settings, a sizable share of responsiveness growth is linked to philanthropic and government-supported research, while portfolio tilt differs sharply across funder types. The contrast between corporate and philanthropic tilt is consistent with market-size–driven innovation in pharmaceuticals (Acemoglu and Linn, 2004) and with time-to-payoff considerations in clinical development (Budish et al., 2015). In policy terms, progress toward burden-aligned research is not automatic: it depends on institutional capacity and on funding structures that sustain attention to diseases with high social burden but weaker market incentives. Philanthropic donors therefore play an outsized role in plugging critical gaps for neglected diseases (World Health Organization, 2024a); sustaining and scaling those investments will be important if the global community is to meet the 2030 Sustainable Development Goals for health. Public agencies and multilateral funders can use burden-adjusted publication metrics to identify under-served diseases and geographies, directing grants or fellowships to places of greatest need, though care must be taken not to reinforce “safe-topic” incentives in evaluation (Wang et al., 2017).

Several caveats apply. Our sample of 308,135 papers results from successive filters (Health Sciences articles, citation and affiliation requirements, identifiable geographic content) applied to 524 JCR-listed journals; access to full texts would yield richer geolocation of study cohorts. OpenAlex’s coverage may undercount regional and non-English-language titles, particularly in parts of Africa and South America. We do not assess methodological quality or treatment effects. Further validation of LLM-based disease tagging for rare conditions would strengthen confidence. These limitations do not undermine the main message: the geography of medical research is shifting toward greater alignment with health needs, yet significant misalignments remain.

An important question for interpretation and policy is whether observed responsiveness reflects net expansion of research or reallocation from other disease–location cells. If gains are largely reallocative, progress in burden alignment may come at the cost of reduced attention elsewhere; if they are expansionary, responsiveness can improve without zero-sum trade-offs. Distinguishing these margins is therefore essential for evaluating the welfare implications of responsiveness and for designing funding regimes that improve both adaptation and equity. Disentangling “capacity deepening” in emerging economies (e.g. growth in China’s, India’s and Brazil’s biomedical infrastructure) from “capacity borrowing” by international consortia and parachute research during major outbreaks (e.g. Ebola 2014, Zika 2016) is a related priority for future work, for example by combining our data with author-affiliation trajectories or emerging North–South collaboration metrics (Nature, 2023).

## Data Availability

All data produced in the present study are available upon reasonable request to the authors

## Acknowledgments

H.Z. acknowledges support from the Accelerate Programme for Scientific Discovery, made possible by a donation from Schmidt Sciences. The authors declare no competing interests.

## Appendix

### A Details on Bibliographic Dataset Construction

#### A.1 Journal Set Identification

To identify journals relevant to this study, we employed the 2023 edition of the *Journal Citation Reports* (JCR), provided by [Clarivate JCR](https://clarivate.com/academia-government/scientific-and-academic-research/research-funding-analytics/journal-citation-reports/). JCR is a widely used journal evaluation tool built on the Web of Science Core Collection, reporting citation-based metrics such as the Journal Impact Factor and assigning each journal to one or more subject categories. These subject categories serve as proxies for disciplinary orientation and are commonly used in bibliometric and research evaluation studies. We linked the journals retrieved from JCR 2023 to [OpenAlex](https://openalex.org/) using both ISSN and eISSN, to further obtain article-level bibliographic data.

We identified 57 medicine-related categories in JCR. Linking these journals to OpenAlex yielded 6,725 journals with unique OpenAlex identifiers. We excluded journals with the word “Review” in the title to omit outlets primarily positioned to publish review articles rather than empirical studies. This reduced the set to 5,861 journals. We then focused on three broad categories that capture general medical research without explicit disease specialization: MEDICINE, GENERAL & INTERNAL, MEDICINE, RESEARCH & EXPERIMENTAL, and PHARMACOLOGY & PHARMACY. This refinement gave us 524 journals, covering 1,065,683 papers indexed in OpenAlex, restricted to documents of type “article” with at least one citation.

#### A.2 Article Corpus Filtering

Starting from 1,065,683 papers, we applied a set of filters to ensure coverage of relevant, attributable, and analyzable research outputs, while also producing a reasonable size of data for our LLM-based retrieval pipeline.

We applied three filters sequentially. First, a domain filter retained papers classified in OpenAlex as only belonging to the Health Sciences domain (741,264 papers; 69.6%). Second, an authorship filter retained papers with valid information on the country or territories of authors’ affiliations (871,934 papers; 81.8%). Third, a language filter retained only papers written in English (1,050,170 papers; 98.5%). Applying all three filters together produced a refined corpus of 603,447 papers (56.6%).

#### A.3 Geographic Context Filtering

To study the geographical dimension of medical research, we further filtered papers to identify those with potential geographical context in their titles or abstracts. Two complementary approaches were used. The GeoText method applied the Python geotext library (https://github.com/elyase/geotext) to extract city and country mentions, identifying 202,790 papers (33.6%). The NER method used a named entity recognition model (dslim/distilbert-NER) to extract LOC and ORG entities as potential location markers, identifying 309,950 papers (51.4%). Retaining papers identified by either method gave a total of 349,737 papers (58.0%).

#### A.4 LLM Retrieval and Linkage

We then processed the 349,737 papers with our LLM-based retrieval pipeline to enrich them with standardized biomedical and contextual information. This step successfully linked 308,135 papers (88.1%) to at least one Medical Subject Heading (MeSH). Among these, 266,532 papers (86.5%) were further linked to at least one level 2 Global Burden of Disease (GBD) category. Funder information was available for 32,057 papers (10.4%), disclosed and retrieved from OpenAlex. The LLM pipeline identified geographical context for 169,262 papers (54.9%).

The resulting curated dataset consists of 308,135 papers, enriched with relevant disease categories, authorship information, funder information, and geographical context.

### B Details on LLM Retrieval of Academic Papers

Our empirical design hinges on two ingredients for every article in our corpus:

1. *paper–data metadata*, which record whether the study uses primary or secondary data, the unit of analysis, temporal coverage, geographic scope, and ownership; and
2. *paper–disease links*, which anchor every disease mention to a controlled biomedical vocabulary so that research effort can be compared with national disease burdens.

We obtain both with a single large-language-model (LLM) call per paper. The model `(gpt-4o-mini-2024-07-18)` receives the article’s title and abstract plus a structured *system prompt* that instructs it to respond with a JSON object. We use the default model hyperparameters.

Strict schema validation is enforced via OpenAI’s structred output format, minimising hallucination and guaranteeing parseability.

#### B.1 LLM Prompt for Paper–Data Metadata Extraction

The prompt casts the model as an expert analyst of biomedical papers, tells it to ignore everything except data-related content, and spells out the required JSON keys (with explicit fall-backs such as NA or empty arrays) to guarantee parseable output.

##### System instructions

We provide the following instructions to the model for each call:

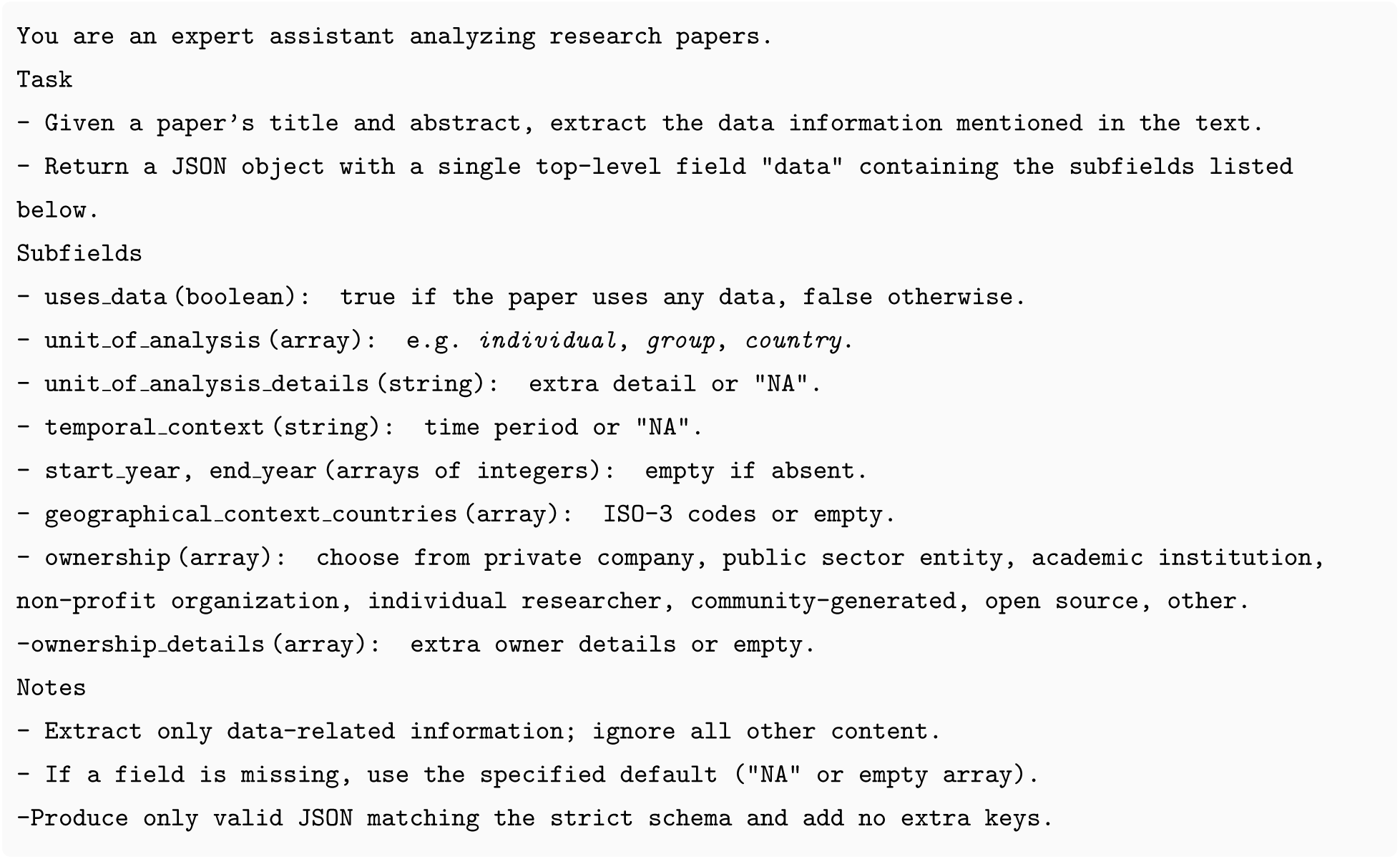

##### User prompt

For each paper, the following message is sent, where {TITLE} and {ABSTRACT} are the raw metadata:

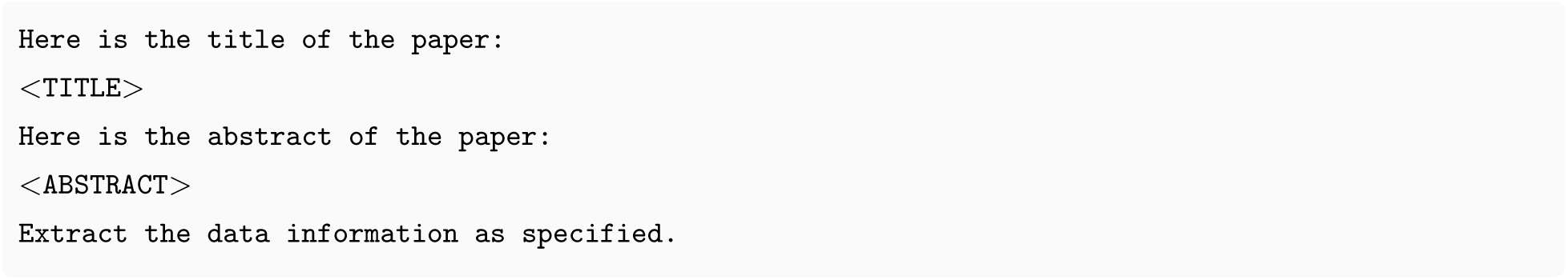

#### B.2 Embedding-based Normalisation to MeSH

All disease strings appearing in the extracted metadata are converted into 1,024-dimensional embeddings using `text-embedding-3-large`. We pre-compute embeddings for the 30,836 preferred

MeSH descriptor terms (including scope notes) and assign each disease the descriptor with the highest cosine similarity.

#### B.3 Limitations

Three caveats merit mention. First, the model works only with titles and abstracts: full-text mining could reveal additional entities but is possible only for open-access papers. Second, assigning each disease mention to a single MeSH descriptor ignores genuine polysemy. Third, information on temporal coverage, comprehensive disease attribution and data ownership is often missing; we therefore carry a dedicated missingness flag into all regressions. None of these constraints undermines the pipeline’s ability to deliver a scalable, reproducible bridge from academic papers to policy-relevant, disease-coded metadata.

### C Details on LLM Retrieval and Structuring of WHO Disease-Outbreak News

We download all Disease Outbreak News (DON) items published on www.who.int/emergencies/disease-outbreak-news. The snapshot used here covers alerts issued between 1996-2025 and yields 3,134 HTML pages. After stripping boiler-plate, we retain four fields per page: the alert title, publication date, canonical URL slug, and full prose.

#### LLM Prompt For Alert Extraction

Each page is passed, one call per alert, to `gpt-4o-mini-2024-07-18` together with a system prompt that instructs the model to return an array of “alerts” containing, for every disease mention, seven slots: disease, geography, date, month, year, cases, deaths, and computed case-fatality ratio if provided. All slots default to NA (not available) when absent. The schema is declared as `“strict“: true` and `“additionalProperties“: false so` that any hallucinated field triggers an automatic retry.

#### System Instructions

We provide the following instructions to the model for each call:

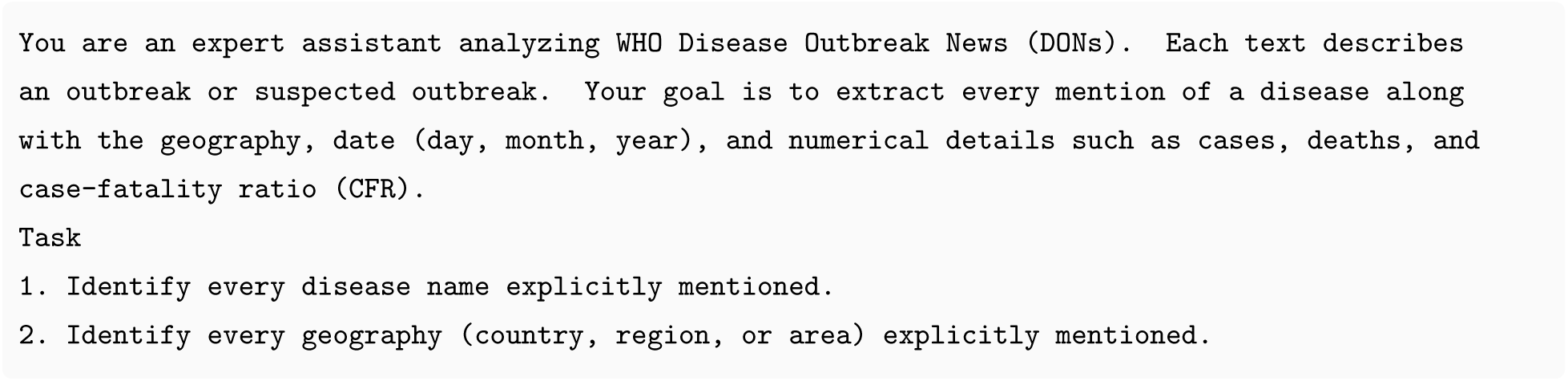
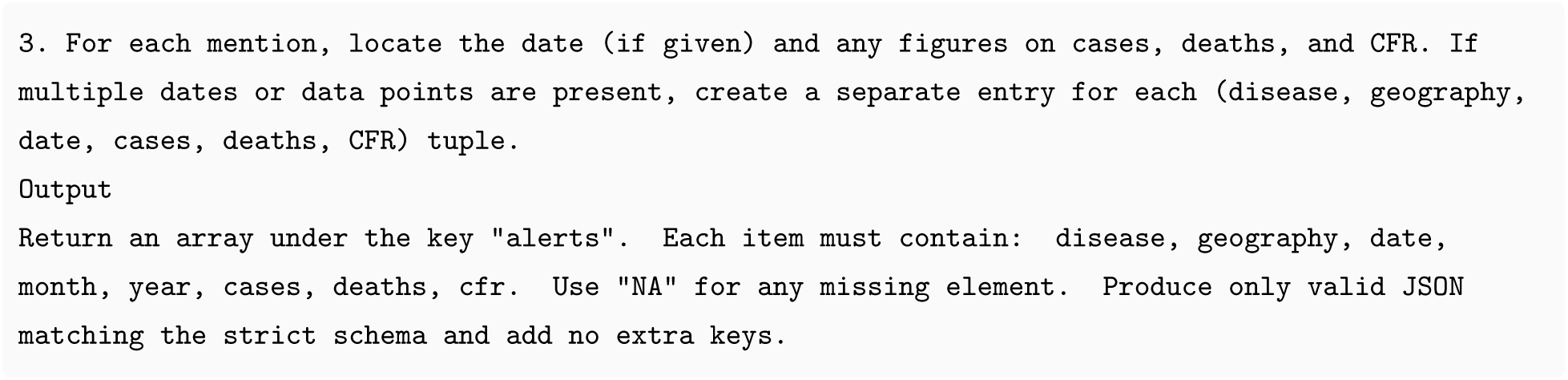

#### User Prompt

For each alert, the following prompt is supplied, where <TITLE>, <DATE>, and <CONTENT> are the page’s metadata and prose:

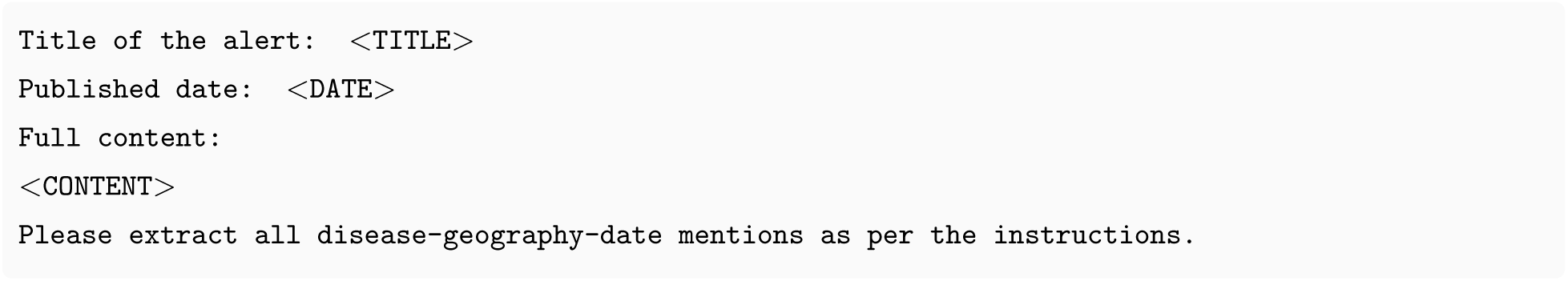

#### Normalising Diseases to MeSH

Free-text disease names are embedded with text-embedding-3-large (1,024 dimensions) and matched to the nearest MeSH descriptor by cosine similarity, exactly as described in Section B.2. The resulting MeSH codes are subsequently linked to ICD-10 and GBD level-2 causes for empirical work.

#### Refining Geography Strings

Alert prose often mixes coarse labels (“*Central Africa*”) with granular places (“*Kasese District*”). To standardise these, we run a second LLM call on the unique set of geography strings (roughly 1,500 after de-duplication) guided by a schema that asks for: city, country (ISO-3), continent (seven-code system), region, and other. Each geography is assigned a deterministic geography identifier, enabling many-to-one joins back to the alert table.

#### System Instructions

We provide the following instructions to the model for each call:

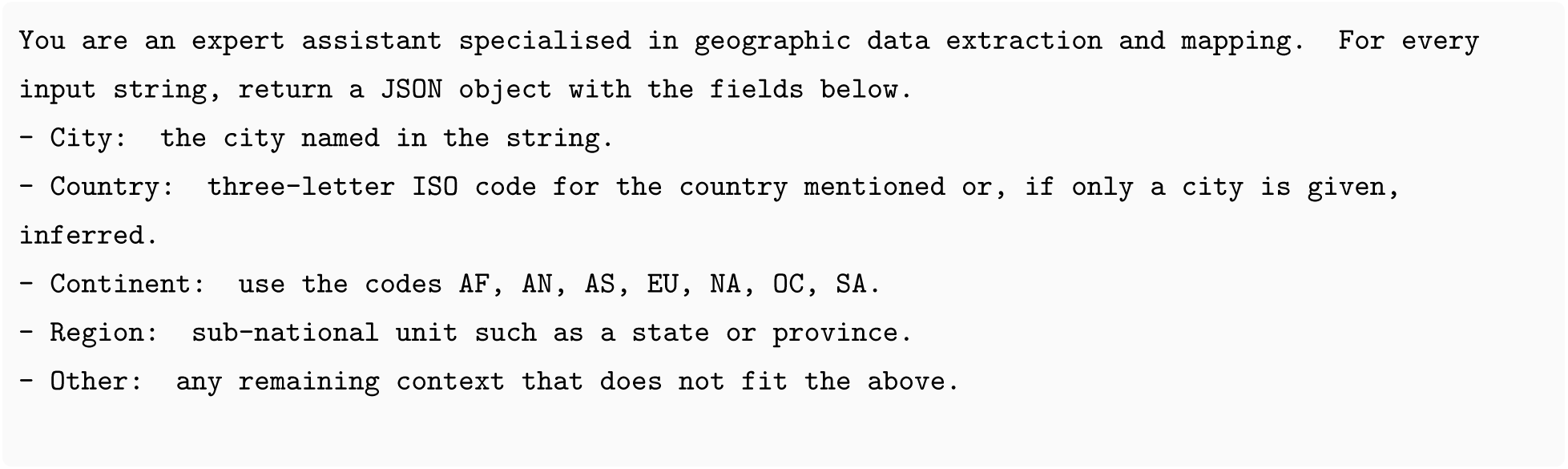
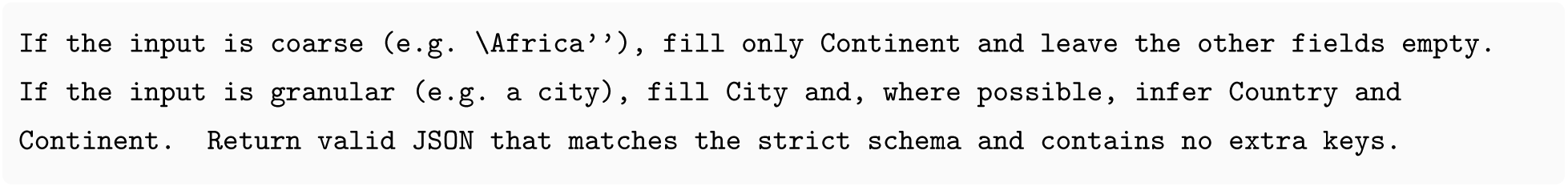

#### User Prompt for Geography

For each distinct geography string we send:

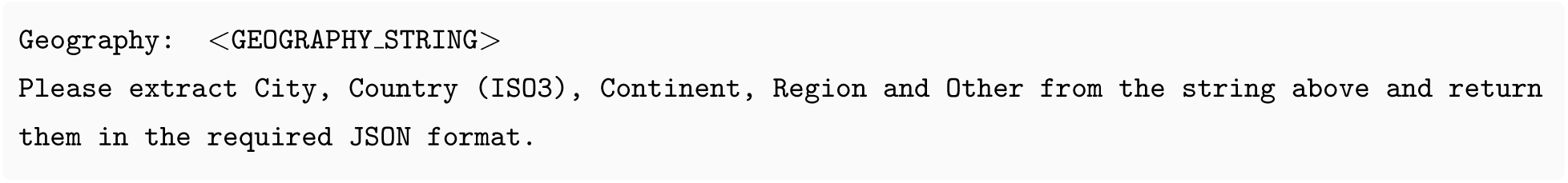

#### Post-processing Steps

1. **Year back-fill.** Where the alert text lacks an explicit year (≈ 6%), we impute it from the publication date.
2. **MeSH tree levels.** A unique MeSH tree cross-walk maps every descriptor to up to three hierarchical levels (mesh tree level 1–3), facilitating aggregation in event-study regressions.

#### Limitations

Three issues deserve note. First, case and death figures follow no common template, so treating every number that precedes “cases” or “deaths” as valid can blur distinctions between suspected and confirmed counts. Second, several DON items cover more than one disease (for instance dengue and chikungunya), and our extraction rightly records each mention but inherits any ambiguity in the source text. Third, where the prose names only a continent, finer geographic slots remain empty, potentially understating sub-national variation. Even so, the pipeline yields the high-frequency, geography-resolved shocks that underpin the event-study design.

### D Details on LLM classification of research funders

We classify every one of the 32,437 funders listed in the 2024 OpenAlex snapshot. Each funder’s display name is sent five times to `gpt-4o-mini-2024-07-18` (temperature 0.7). The model is instructed to return a JSON object with six Boolean keys: direct government, independent public, private corporate, private philanthropic, academic institution, and hybrid. Strict schema validation `(“strict”: true, “additionalProperties”: false)` guarantees parseable output.

#### System Instructions

We provide the following instructions to the model for each call:

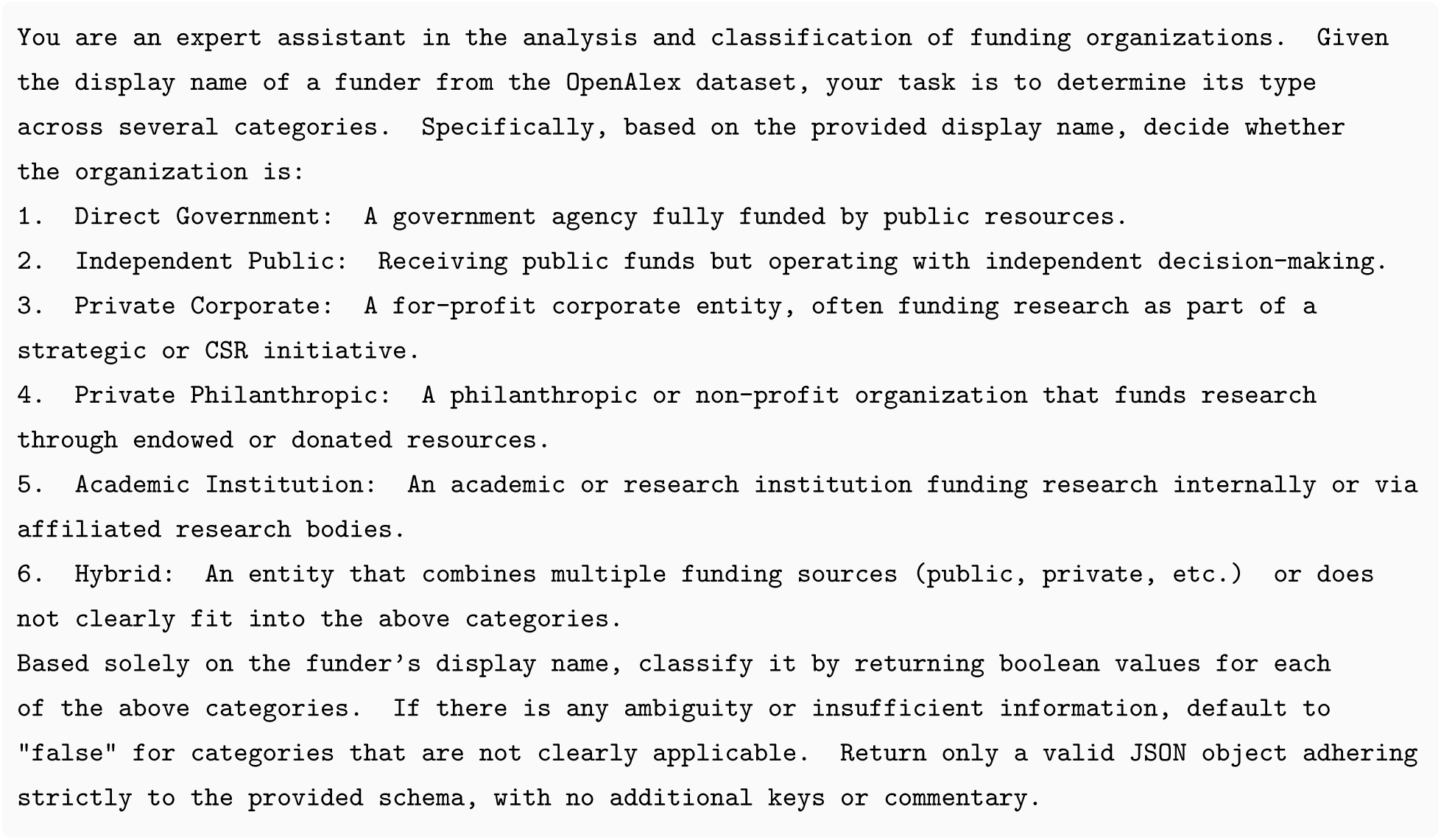

#### User Prompt

For each funder, the following prompt is supplied, where “<DISPLAY NAM>” is the name of the funder as provided by OpenAlex:

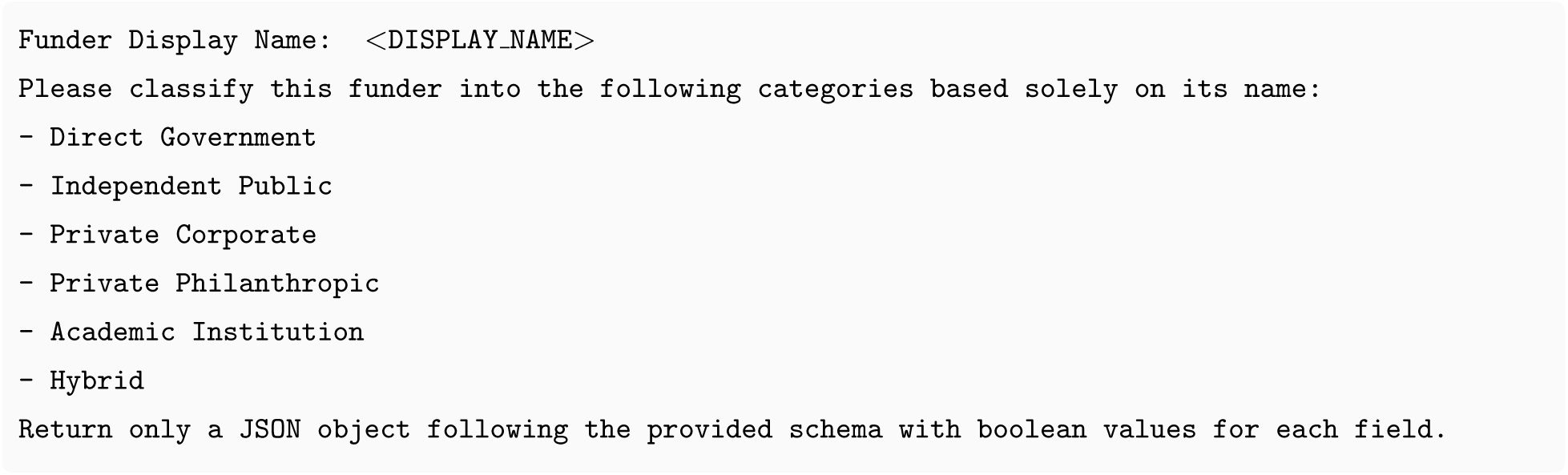

#### Aggregation and Consistency

For each (*id*, category) pair we take the modal value across the five responses; ties are broken at random. Across the six categories, 96–99% of funders receive identical labels in all five calls, indicating high internal reproducibility despite the stochastic sampling (model temperature was set at 0.7).

#### Category Prevalence

In the universe of funders, 36% are labelled academic institution, 34% private philanthropic, 17% direct government, 9% private corporate, 4% independent public, and 2% hybrid. Because the categories are not mutually exclusive, a single funder can carry multiple true flags.

#### Potential Limitations

Classification relies on the name alone, which can be ambiguous or language-specific; mixed-funding bodies may fit more than one bucket; and the six categories are an imperfect proxy for more granular legal forms. For our purposes (broadly contrasting public, corporate, philanthropic and hybrid sources) these coarse buckets are adequate, and the high agreement rate suggests that any residual misclassification is unlikely to change the qualitative results.

## Footnotes

1 UNESCO’s 2023 Science Report estimates that Africa, with roughly 12.5% of the world’s population, accounts for under 1% of global research output and invests barely 0.5% of GDP in R&D—ten times below high-income averages (Lewis et al., 2021).

2 Recent syntheses highlight the gap: the Access to Medicine Foundation’s 2024 Index finds that most large pharmaceutical firms “are still not maximising their potential to reach patients in LMICs” (Access to Medicine Foundation, 2024); a Lancet Viewpoint on global R&D priorities highlights the same tension for non-communicable diseases (Vollset et al., 2024); and WHO’s 2024 NTD report stresses that R&D bottlenecks remain a core barrier to disease elimination (World Health Organization, 2024a).

3 See also (Barakati et al., 2025), (Garg and Fetzer, 2025) for examples where LLMs are used to recover structured scientific relationships from text, illustrating the broader potential of LLM-assisted measurement.

## Notes

### Competing Interest Statement

The authors have declared no competing interest.

### Funding Statement

Zhou is supported by funding from the Accelerate Programme for Scientific Discovery, made possible by a donation from Schmidt Sciences.

### Summary of Updates

This version revises the title, abstract, and wording throughout the manuscript to improve clarity and framing. There are no changes to the data, empirical analysis, methods, results, figures, or tables. The substantive findings and conclusions remain unchanged.

